# Micro-Costing Analysis for Human Papillomavirus (HPV) Vaccination Services for Out-of-School (OOS) Girls in Indonesia: Evidence from a Pilot Study

**DOI:** 10.64898/2026.06.24.26356497

**Authors:** Adiatma Yudistira Manogar Siregar, Indah Amelia, Rahma, Priscillya Hotma, Faisal Madjid Alyasa, Lili Nur Indah Sari, Shabrina Mumtazah, Rizki Andini, Niken Widyastuti, Tetrawindu Hidayatullah, Atiek Anartati, Sonali Patel, Devi Anisiska, Endang Budi Hastuti, Prima Yosephine

## Abstract

Cervical cancer remains a major public health challenge in Indonesia, with Human Papillomavirus (HPV) infection responsible for nearly all cases. The government has integrated HPV vaccination into the national School Children Immunization Month (BIAS). However, out-of-school (OOS) girls remain difficult to reach because they are not covered by the school-based vaccination platform. Micro-costing approach was used to estimate both economic and financial costs of delivering HPV vaccination to OOS girls in Bekasi City, Bandar Lampung City, and Bangka District. This study aimed to estimate the cost of delivering a single-dose HPV vaccination program to OOS girls and to project the national-level cost of scaling up the program. Costs were categorized into outreach efforts, such as community mobilization and identification of girls, and delivery components including logistics, storage, and administration. A scale-up costing analysis was also conducted to estimate national-level costs. The results show that the cost per vaccinated OOS girl aged 11 ranged from US$ 22.77 to 38.55, while the financial cost range from US$ 12.70 to 29.50, varied by implementation context, with outreach activities, transportation, and vaccine delivery supplies identified as the main cost drivers. The estimated national annual economic and financial cost of scaling-up HPV vaccination for OOS girls are US$ 644,215.23 and US$ 483,138.35, respectively. Our national cost estimates show that reaching and vaccinating OOS girls accounts for less than 3% of the national immunization budget. Factors such as local context and implementation challenges should be recognized as they may directly influence the cost and hinder the program scale-up.

## Introduction

Cervical cancer remains one of the major public health challenges in Indonesia, Indonesia ranks third globally in the number of cervical cancer cases compared to other countries [1]. The disease accounts for approximately 40,000 new cases and 20,000 deaths annually. Nearly 99.7% of cervical cancer cases are caused by the Human Papillomavirus (HPV) [2]. Although HPV can also cause other types of cancers in both women and men, cervical cancer remains the most common cancer associated with HPV infection [3].

In response to these challenges, Indonesia has undertaken several steps, including strengthening preventive efforts through HPV immunization and improving screening methods. Through the National Action Plan (RAN) for Cervical Cancer Elimination 2023–2030 [4], the government has set a target of achieving 90% HPV immunization coverage among girls by the age of 15. Implementation of the free HPV vaccination program is integrated into the annual School Children Immunization Month (BIAS), alongside Measles-Rubella (MR) and Tetanus and Diphtheria (DTTD) immunizations.

However, significant challenges remain in achieving equitable HPV immunization coverage. A study in Indonesia highlights persistent disparities in coverage, particularly in ensuring that all girls complete the vaccination series [5]. Recent data from Ministry of Health (MoH), HPV vaccination coverage in Indonesia reached 91.1% in 2025 after the implementation of the single-dose since 2025 [6].

One of the factors contributing to challenges in achieving 100% coverage is the difficulty in reaching out-of-school (OOS) girls, who are not accessible through the formal school-based BIAS mechanism. Current HPV vaccination efforts in Indonesia are predominantly school based through the *Bulan Imunisasi Anak Sekolah* (BIAS) mechanism; however, this system lacks a specific national strategy to reach those outsides of formal settings. Consequently, OOS girls remain underserved. Based on the OOS estimation in this study (Table 3), the estimated number of OOS girls aged in Indonesia is approximately 25,354 nationally. Many of these girls come from marginalized backgrounds, limiting their access to HPV vaccination. Additionally, approximately 12,953 educational institutions, representing 2.34% of all educational institutions in Indonesia, operate as non-formal schools, such as Community Learning Centers (PKBM), Islamic boarding schools, and other alternative education pathways [7].

In Indonesia reaching OOS girls presents challenges related to cultural norms, misinformation, and concerns among some parents regarding HPV vaccination [8]. As HPV vaccination is primarily delivered through schools, religiously affiliated and non-formal education settings may require more intensive engagement, as previous evidence suggests that faith-based school communities may be more hesitant to encourage HPV vaccination. This is also supported by a study in Indonesia showing that the halal–haram status of the HPV vaccine has become increasingly crucial, indicating that religion plays an important role in health decision-making [9,10]. In addition, parents and teachers may have limited confidence and knowledge to discuss sexual and reproductive health topics with young people, which can affect communication and acceptance of HPV vaccination[11].

Similar challenges are also observed in low and middle-income countries, which require additional community-based strategies and proactive sensitization efforts [12,13]. The situation is further compounded by socioeconomic vulnerabilities, which many OOS girls live in marginalized communities, engage in informal labor [14], face early marriage, or lack supportive caregiving structures [15]. To respond to these barriers, multiple strategies have been introduced in Bangladesh, Burkina Faso, Ethiopia, The Gambia, Mali, and Senegal including cross-sectoral coordination platforms, stakeholder engagement, and the dissemination of inclusive Communication, Information, and Education (CIE) materials [12,14].

Subsequently, reaching OOS girls may require additional resources for identification, mobilization, outreach, and the provision of vaccination services, as demonstrated by implementation experiences in Mozambique, Zimbabwe, Peru, Uganda, Vietnam, the UK, Belgium, and several U.S. federal programs [16]. Consequently, the cost per vaccinated OOS girl is estimated to be significantly higher than the cost per vaccinated girl under school-based programs [16–18]. A systematic cost analysis is therefore essential to assess cost per vaccinated OOS girls to ensure the sustainability and effectiveness of HPV vaccination programs, especially for OOS groups who are epidemiologically and socially more vulnerable to gaps in routine health interventions [16]. However, comprehensive cost estimates for Indonesia are currently limited, if any.

This study aims to estimate the cost of delivering a single-dose HPV vaccination program to OOS girls and to project the national-level cost of scaling up the program. The analysis includes mapping outreach processes and identifying cost components at multiple levels of the health system and community. The findings are expected to provide evidence-based input for policy makers to design efficient financing strategies that ensure equal rights to preventive care for all girls. By expanding HPV vaccination coverage among OOS populations, the government can not only advance toward national cervical cancer elimination targets under the 2023–2030 National Action Plan but also mitigate substantial future treatment costs associated with the disease.

The costing is based on an HPV vaccination pilot project targeting OOS girls in Indonesia, implemented by Clinton Health Access Initiative (CHAI) from October to December 2023. The pilot included target identification, outreach, and evaluation conducted in sequential stages at the city and community levels [19,20]. The pilot was carried out in Bekasi City, Bandar Lampung City, and Bangka District, three urban settings with relatively high HPV vaccination coverage in 2024: 86%, 93%, and 88%, respectively. [19,20]. This pilot emphasized strengthening cross-sectoral coordination and community-based outreach strategies to effectively reach 11-year-old girls who are not enrolled in formal education (i.e. either out of school completely or undergoing informal education) [19]. The pilot was initiated by Clinton Health Access Initiative (CHAI) and involved a wide range of cross-sector stakeholders.

## Methods

### Study design and overview

The study employed micro-costing analysis to estimate the cost of HPV vaccination outreach and service delivery for OOS girls in Bekasi City, Bandar Lampung City and Bangka District. The analysis was conducted from both financial and economic perspectives, capturing direct program expenditures as well as the costs associated with personnel time, equipment, and facility use.

The costing framework covered the outreach and full-service delivery pathway, including target identification and coordination, target validation, stakeholder socialization, and vaccination implementation. OOS girls were defined as 11-year-old girls who were either not enrolled in formal education, attending informal or non-formal educational institutions, or completely out of school.

Cost data were collected through document review, field observation, and in-depth interviews with key stakeholders, including district/city health offices, primary health centers (*puskesmas*), and partner institutions involved in HPV vaccine delivery. In addition to site-level costing, the study developed a national scale-up simulation to estimate the potential cost of expanding a single-dose HPV vaccination strategy for OOS girls across Indonesia.

### Definition of OOS girls

Following the intervention by CHAI, we defined OOS girls as those who are not enrolled in formal education, including two main groups. First, those enrolled in informal or non-formal education (i.e., attending non-formal learning settings such as Islamic boarding schools/*pesantren*, homeschooling, and other similar programs). Second, those who are completely out of school (i.e., not attending any educational institution), which includes those residing in non-educational institutions (e.g., orphanages, rehabilitation centers, and similar facilities) as well as those living in the community outside any institutional setting [21]. We limited our analysis to OOS girls aged 11 years.

### Study setting and implementation

#### Study implementation

This study employs a purposive sampling method to select relevant stakeholders from CHAI, district/city health offices (DHO), primary health centers (*puskesmas*), and other stakeholders perform interviews related to costing analysis. The selection criteria were adjusted based on discussion with DHO and CHAI or through a snowball sampling approach during data collection (Table 7).

#### HPV vaccination delivery activities

The activities were based on the pilot initiated by CHAI in 2023. The pilot implementation involved a wide range of cross-sector stakeholders, including City/District Health Offices, *Puskesmas*, Education Offices, the Ministry of Religious Affairs, Social Affairs Offices, Civil Registration Offices, community organizations, and CHAI as the main facilitator [20]. CHAI played a central role in facilitating cross-sector coordination, developing strategies, and documenting lessons learned to support the integration of OOS approaches into the national immunization program [19,20]. The activities are grouped into four main groups, namely target identification and coordination, target validation, socialization to involved stakeholders, and HPV vaccination (table 1).

**Table 1.** Identification of HPV vaccine delivery activities for out-of-school (OOS) girls.

| Process | Activities | Organizer | Participants |
| --- | --- | --- | --- |
| Identification and coordination | Identification of cross-sectoral roles and responsibilities | District/city health office | *Puskesmas*, education office, social affairs office, ministry of religious affairs, and other relevant stakeholders. |
|  | Quantification, identification, and categorization of OOS girls | District/city health office | *Puskesmas*, education office, social affairs office, ministry of religious affairs, and other relevant stakeholders. |
|  | Target identification, outreach planning, and logistics requirement estimation | District/city health office | *Puskesmas*, education office, social affairs office, ministry of religious affairs, and other relevant stakeholders. |
| Target validation | Target validation: Communication via whatsapp | *Puskesmas* | *Puskesmas* and nonformal school. |
|  | Target validation: School visits | *Puskesmas* | *Puskesmas* and nonformal school. |
|  | Validation of targets from marginalized groups | *Puskesmas* | *Puskesmas* and cadre. |
| Stakeholder socialization | Socialization with nonformal schools: School visits | *Puskesmas* | *Puskesmas*, cadre and nonformal schools |
|  | Socialization with nonformal schools at the *puskesmas* | *Puskesmas* | *Puskesmas*, cadre, nonformal schools and other relevant stakeholders. |
| Vaccination implementation | Implementation of HPV immunization | *Puskesmas* | *Puskesmas* and nonformal school. |

CHAI’s financing for this project at the district/city level for coordination process was calibrated to match the existing routine activity budgets of the relevant offices to ensure sustainability and institutional alignment, as the activities supported by CHAI were considered an integral part of each stakeholder’s established routine activities rather than standalone interventions. In addition, staff working time allocated to CHAI-supported activities was also captured in this calculation, though measured from the perspective of the employees themselves. The routine health office meetings with cross-sectoral stakeholders conducted four times per year, with the presentation on the School Children Immunization Month (BIAS) activities for HPV vaccination delivered once a year, covering socialization and data collection on the target population of OOS girls. This implementation scheme was applied across all study locations.

### Costing framework

#### Costing approach

We used micro-costing, complemented by document review and in-depth interviews, to provide contextual insights regarding the cost data and their background. The framework (figure 1) begins by identifying key inputs (e.g. human and financial resources) utilized in the delivery process to deliver a single dose HPV vaccination program, encompassing logistics, vaccine storage, transportation, administration, as well as information, education, and communication (IEC) activities. It also includes the identification and validation of OOS girls and community mobilization efforts. The use of resources and associated expenditure related to these activities are systematically assessed through cost analysis to determine both recurrent and capital costs. We calculated the total cost of each stakeholder in relation to respective activities, as shown in table 1.

**Figure 1.**
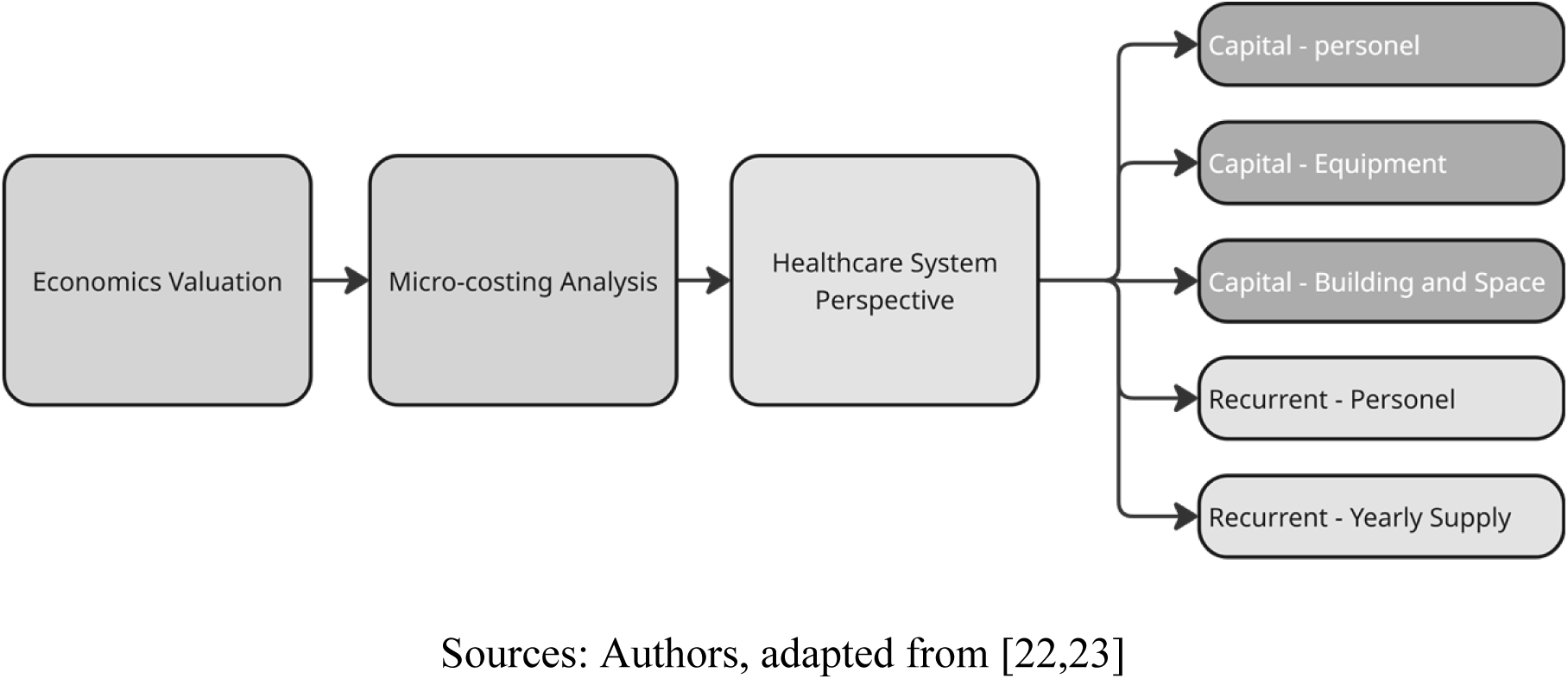
*Micro-costing* Framework

Capital costs encompass expenditures associated with health workforce training and capacity development, equipment, and buildings. Recurrent costs include the value of healthcare workers’ time devoted to program activities and annual needs for service delivery (e.g. vaccines, syringes, and safety boxes). By combining capital and recurrent cost components, this study calculates the program’s total expenditure and subsequently derives the unit cost per OOS girl fully vaccinated against HPV.

### Micro-costing components and assumption

The summary of assumptions, cost components, and data sources are presented in table 2, and we aligned our approach with the CHEERS checklist (Table 8) [24], thus our costing method is comparable to other studies. We present our results both as financial and economics costs. The financial costs only encompass costs related to items that are utilized specifically for the implementation (e.g. consumable supplies, transportation), while economic costs encompass all items included in financial costs as well as other costs related to the implementation from health system perspective (e.g. wage, building).

**Table 2.**
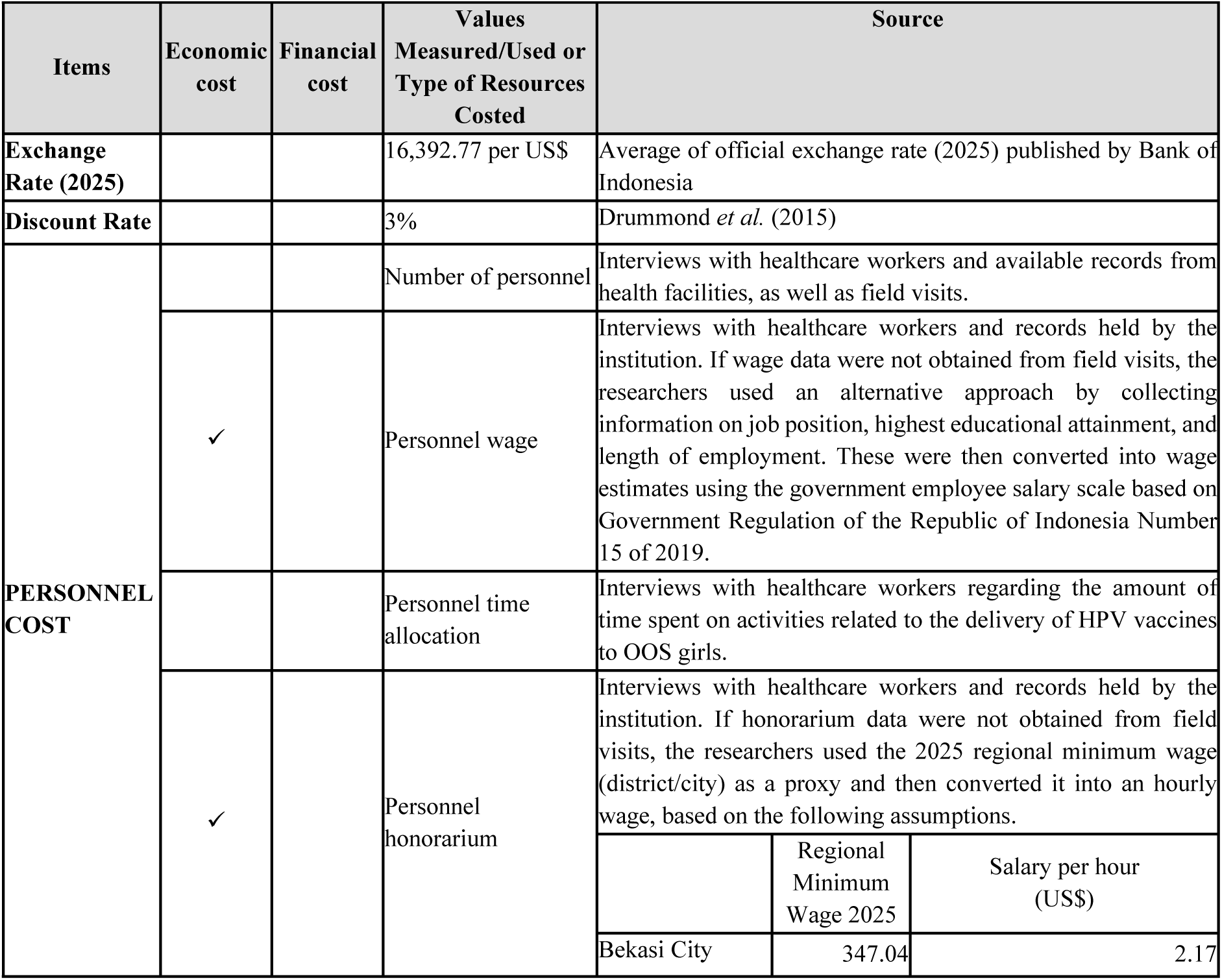
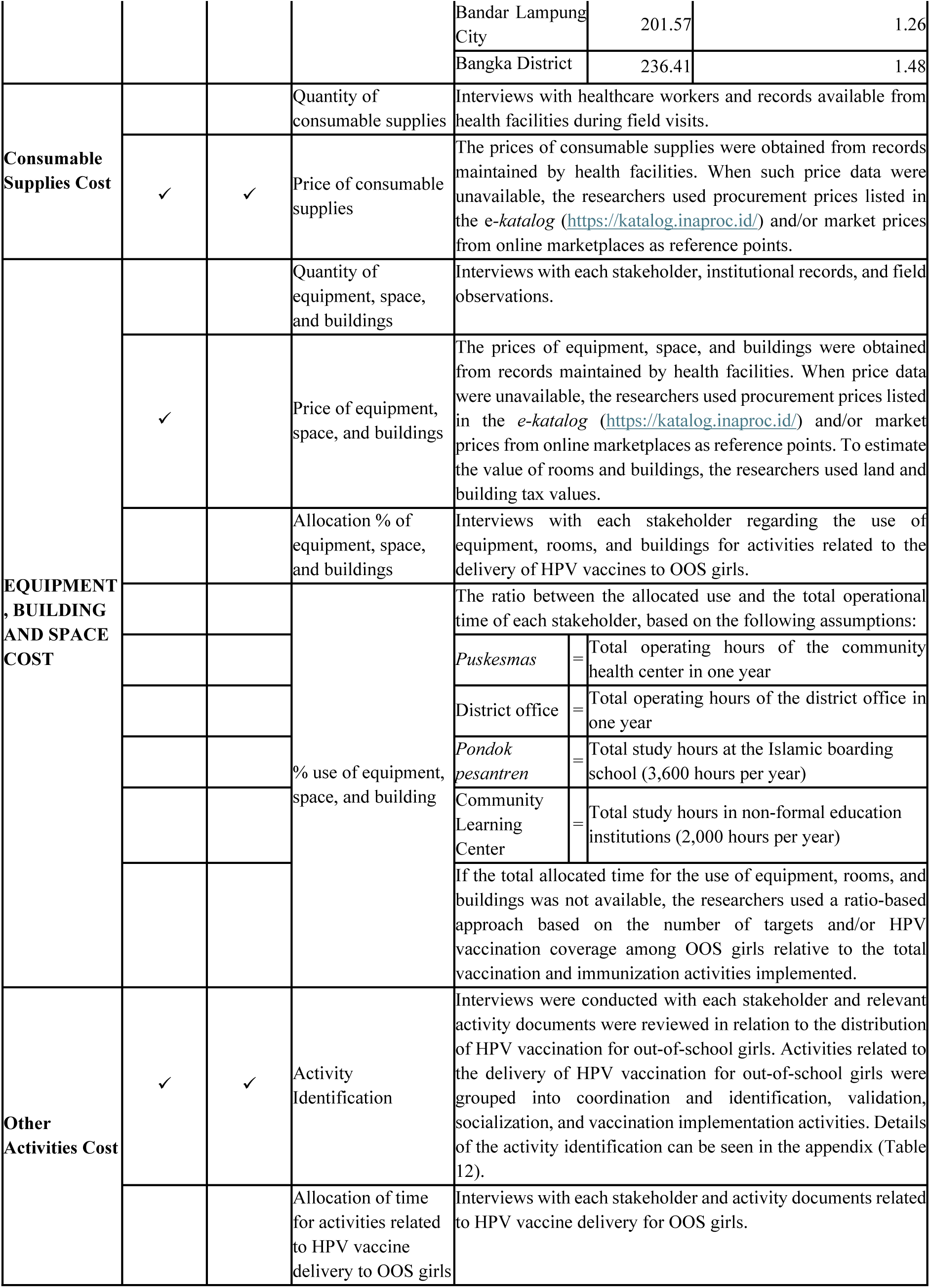
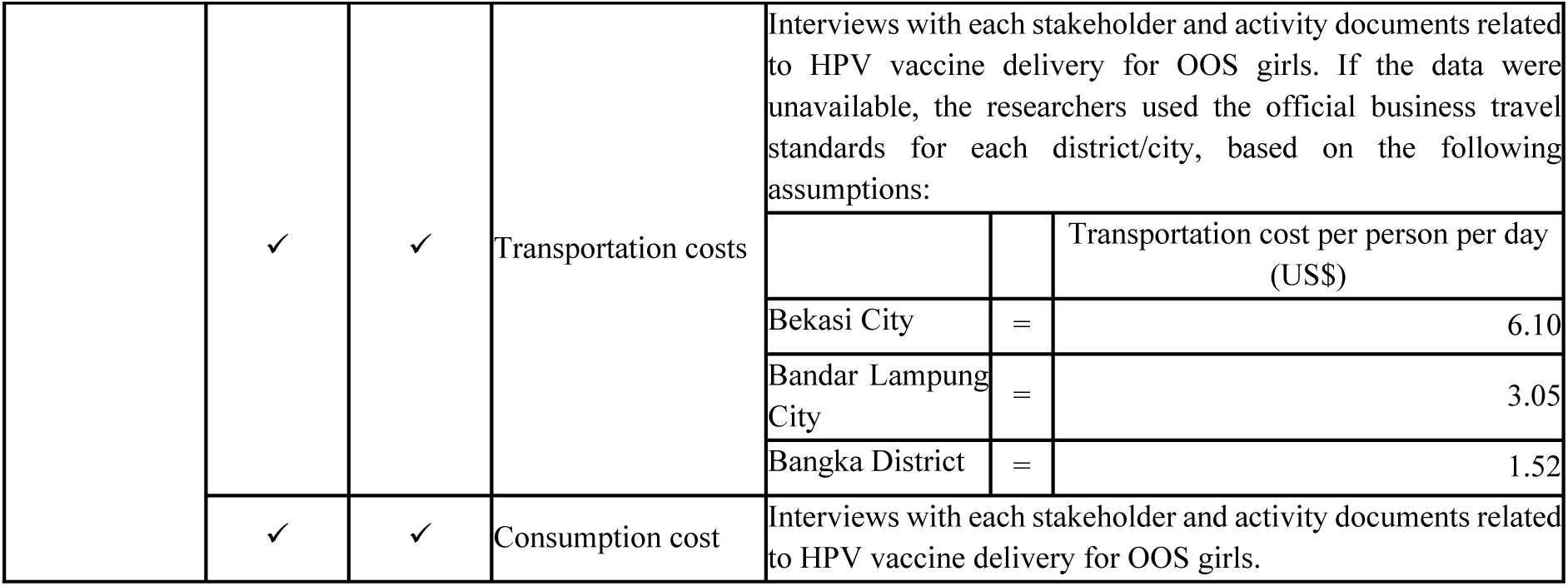
Assumption and Values Used on Micro-costing Analysis.

### Scale-up simulation strategy

The scale-up simulation was developed to estimate the costs of a single-dose vaccination strategy specifically for OOS girls when expanded up to the national level. First, we mapped each province into three categories representing similarities in characteristics and implementation needs with the three study areas. This mapping determined the reference cost used for each province based on the corresponding study area, thereby ensuring that the costing approach remained consistent with the context of the study sites used as benchmarks. Second, we adjusted the reference unit costs using a regional remuneration adjustment coefficient, namely the 2025 INKINDO Index, to capture differences in cost levels across provinces. Finally, the unit cost of outreach and delivery for each province was multiplied by the target population of OOS girls aged 11, as explained in Table 3. The resulting costs for each age group were then summed to obtain the total national scale-up cost. The assumption for scale up estimation is presented in table 3. either fully out of school + those who undergo informal schooling

**Table 3.** Assumption and Values Used on Scale-up Estimation.

| Item | Values Measured/Used or Types of Resources Accounted For | Source |
| --- | --- | --- |
| Total Cost | Cost of HPV vaccination outreach for OOS girls per target | A micro-costing estimate conducted at *puskesmas* level, presented in detail (US\$), using the target population for HPV vaccination among OOS girls as the denominator. |
| | Cost of HPV vaccine delivery for OOS girls per target | A micro-costing estimate conducted at *puskesmas* level, presented in detail (US\$), using successfully vaccinated OOS girls as the denominator. |
| Adjustment Coefficient | Adjustment coefficient | The 2025 Provincial Standard Remuneration Index. This index was used as a cost adjustment coefficient across regions to adjust the cost estimates obtained from the study sites to reflect differences in labor cost levels in other provinces. By using this standard remuneration index, the scale-up cost estimates are expected to better capture variations in operational cost conditions across regions in Indonesia (INKINDO, 2025). |
| Target of OOS girls | Number of OOS girls aged 11 years | The number of OOS girls was estimated by combining school participation rates (APS) with population data. The calculation followed the formula: $OOS = (1 - APS) \times N_{11}$. APS data were obtained from Statistics Indonesia (BPS) publications for 2025, representing the proportion of children attending school within the relevant age group[25]. Meanwhile, the population of 11-year-old girls ($N_{11}$) was derived from the 2020 Population Census (BPS)[26]. This method provides an estimate of the number of girls who are not enrolled in school within each province. |

### Ethical considerations

This study received ethical approval from the Research Ethics Committee of Universitas Padjadjaran under approval number 1025/UN6.KEP/EC/2025. All data collection procedures were conducted in accordance with relevant ethical guidelines and regulations. Participation in interviews and data collection activities was voluntary, and informed consent was obtained from all respondents prior to data collection. Confidentiality and anonymity of participants and institutional information were maintained throughout the study.

## Results

Based on the results of the micro-costing estimation conducted at the district/city level (Table 4), the three study sites showed relatively small variations in total costs. However, when analyzed based on the unit cost at each Community Health Center, greater cost variation was observed, given that each district/city has a different number of Community Health Centers. Among the three sites, Bekasi City had the lowest unit cost at US$ 2.56.

**Table 4.** Estimated Financial and Economic Cost of HPV Immunization for OOS Girls at the District/City Level (US$)

|  | Bekasi City |  | Bandar Lampung City |  | Bangka District |  |
| --- | --- | --- | --- | --- | --- | --- |
|  | Financial (%) | Economic (%) | Financial (%) | Economic (%) | Financial (%) | Economic (%) |
| District health office | 12.20<br>(100%) | 82.33<br>(60.75%) | 15.44<br>(100%) | 77.34<br>(59.58%) | 71.48<br>(100%) | 99.33<br>(72.59%) |
| Others district government office* | - | 53.13<br>(39.22%) | - | 52.47<br>(40.42%) | - | 37.50<br>(27.41%) |
| **Total cost** | **12.20** | **135.46** | **15.44** | **129.81** | **71.48** | **136.83** |
| Number of *puskesmas* | 53 | 53 | 15 | 15 | 12 | 12 |
| *Unit cost* | *0.23* | *2.56* | *1.03* | *8.65* | *5.96* | *11.40* |

Source : Authors' calculation

Based on the type of activity, target validation and socialization conducted by *puskesmas* through school visits were activities implemented in all study areas for this program. Meanwhile, online socialization with schools was conducted only in Bekasi City. In addition, the calculations presented in Table 4 also included the estimated cost per *puskesmas* for activities carried out at the district/city health office level. Bekasi City emerged as the study location with the highest total cost for outreach and HPV immunization implementation. When calculated per target, the outreach and immunization cost per child in Bekasi was relatively high, amounting US$ 38.55 per vaccinated target. In-depth interviews suggested that the higher unit cost observed in Bekasi was associated with the labor-intensive process of identifying and verifying eligible OOS targets within a highly dynamic urban setting. Informants reported that non-formal institutions were often unstable or no longer operational, requiring repeated physical verification and extensive data matching, which increased time and personnel requirements. Additional operational inputs were also reported when reaching marginalized populations, including the involvement of multi-sectoral outreach teams and higher travel allowances due to security considerations.

**Table 5.** Estimated Economic Cost of HPV Immunization for Out-of-School (OOS) Girls (US$)

| Activities | Bekasi City | Bandar Lampung City | Bangka District |
| --- | --- | --- | --- |
| Target identification, outreach planning, and logistics requirement estimation | 239.01<br>(18.79%) | 42.52<br>(3.31%) | 23.31<br>(7.88%) |
| Target validation: Communication via *whatsapp* | 3.41<br>(0.27%) | 7.07<br>(0.55%) | 13.88<br>(4.69%) |
| Target validation: School visits | 53.69<br>(4.22%) | 74.44<br>(5.80%) | 22.56<br>(7.62%) |
| Socialization with nonformal schools at the *puskesmas* | 181.65<br>(14.28%) | 67.67<br>(5.28%) | - |
| Socialization with nonformal schools: School visits | 70.54<br>(5.55%) | 50.92<br>(3.97%) | 52.85<br>(17.86%) |
| Socialization with nonformal schools: Online | 2.01<br>(0.16%) | - | - |
| Validation of targets from marginalized groups | 42.63<br>(3.35%) | 43.27<br>(3.37%) | - |
| Cost of activities at the district/city office level (table 6) | 2.56<br>(0.20%) | 8.65<br>(0.67%) | 11.40<br>(3.85%) |
| **Total outreach cost** | **595.49**<br>**(46.81%)** | **294.55**<br>**(22.97%)** | **124.00**<br>**(41.90%)** |
| Number of outreach targets | 79 | 73 | 54 |
| *Outreach cost per target* | 7.54 | 4.03 | 2.30 |
| **Total cost of HPV vaccination: Single Dose** | **676.64**<br>**(53.19%)** | **988.00**<br>**(77.03%)** | **171.96**<br>**(58.10%)** |
| Number of targets successfully vaccinated | 33 | 56 | 13 |
| *HPV vaccination cost per target successfully vaccinated* | 20.50 | 17.64 | 13.23 |
| **Total outreach and HPV vaccination cost** | **1,272.13** | **1,282.55** | **295.96** |
| *Outreach and HPV vaccination cost per target successfully vaccinated* | 38.55 | 22.90 | 22.77 |

Note : The percentage for each cost component is calculated by dividing the component cost by the total outreach and HPV vaccination cost.
Source : Authors' calculation

### Cost structure and main drivers

We found that the economic cost of the HPV vaccination program for OOS girls was driven largely by routine operational expenditures, such as labor costs, consumable supplies, and outreach activities, rather than by major one-time capital expenditures (for example, equipment purchases). Labor costs consistently represented the largest recurrent cost component, reflecting the labor-intensive nature of the program in identifying, mobilizing, and following up OOS girls. In this study, labor costs are treated as actual expenditures rather than opportunity costs, given that the program resulted in a heightened workload for healthcare personnel. Findings from in-depth interviews indicated that personnel time was heavily utilized for target identification, data validation, and cross-sectoral coordination activities. In addition, transportation costs also constitute an important component of the program, particularly for outreach activities requiring field visits and community engagement. Training costs did not appear because vaccination services for OOS girls followed the same HPV vaccination procedures as those applied in formal schools, therefore did not require additional training for health workers. Using the financial costs results, the cost of outreach is mostly dominated by, among others, transportation and consumption costs. On the other note, the cost of HPV vaccination for OOS girls is dominated by supplies costs (e.g. vaccine) and vaccination related activities (e.g. target identification, validation, and outreach).

### Scale-up cost of HPV vaccination for OOS girls

The scale-up results below present estimates of the costs required for outreach and delivery of HPV vaccination for OOS girls in each province, based on the HPV vaccination costs for OOS girls observed in Bekasi City, Bandar Lampung City, and Bangka District.

Table 6 presents the estimated provincial scale-up cost of HPV vaccination for OOS girls in Indonesia. The table summarizes the total costs required for activities related to outreach and vaccine delivery. These scale-up costs represent the main operational components required to support HPV vaccination outreach for OOS girls across provinces. Approximately 18.50% is allocated to outreach activities, which include efforts to identify, mobilize, and engage OOS girls across provinces. The remaining 81.50% is dedicated to vaccine delivery, covering the logistical and operational components necessary to administer the HPV vaccine. At the national level, the total estimated vaccine delivery economic cost for OOS girls was US$ 644,215.23 per year (median US$ 7,464.37, IQR 25% US$ 3,260.89 – IQR 75% US$ 14,564.13) or US$ 38.56 per fully vaccinated OOS girl. Using financial cost, the total estimated vaccine delivery for OOS girls US$ 483,138.35 per year (median US$ 5,556.42, IQR 25% US$ 2,198.08 – IQR 75% US$ 11,548.04) or US$ 28.92 per fully vaccinated OOS girl.

**Table 6.** Estimated Scale-Up Financial and Economic Cost of HPV Vaccination Outreach for Out-of-School Children by Province.

| Province | Total Outreach Cost |  | Total Full Pathway Cost |  |
| --- | --- | --- | --- | --- |
|  | Financial | Economic | Financial | Economic |
| Aceh | 723.58 | 1,319.52 | 5,821.21 | 7,489.85 |
| North Sumatra | 1,508.69 | 2,751.25 | 12,137.42 | 15,616.59 |
| West Sumatra | 406.09 | 740.55 | 3,267.01 | 4,203.49 |
| Riau | 28.22 | 388.61 | 2,149.23 | 3,852.73 |
| Jambi | 30.79 | 423.93 | 2,344.62 | 4,202.98 |
| South Sumatra | 945.09 | 1,723.46 | 7,603.21 | 9,782.66 |
| Bengkulu | 300.26 | 547.56 | 2,415.60 | 3,108.03 |
| Lampung | 718.66 | 1,310.55 | 5,781.61 | 7,438.90 |
| Bangka Belitung Islands | 10.63 | 146.36 | 809.45 | 1,451.03 |
|  | Financial | Economic | Financial | Economic |
| Riau Islands | 132.90 | 242.36 | 1,069.20 | 1,375.69 |
| Special Capital Region of Jakarta | 3,517.19 | 4,645.52 | 18,182.08 | 23,757.74 |
| West Java | 12,572.95 | 16,606.42 | 64,995.72 | 84,927.09 |
| Central Java | 8,179.83 | 10,803.96 | 42,285.53 | 55,252.67 |
| Special Region of Yogyakarta | 646.84 | 854.35 | 3,343.83 | 4,369.24 |
| East Java | 13,866.63 | 18,315.12 | 71,683.38 | 93,665.57 |
| Banten | 3,315.06 | 4,378.54 | 17,137.13 | 22,392.35 |
| Bali | 835.50 | 1,103.53 | 4,319.12 | 5,643.60 |
| West Nusa Tenggara | 2,156.13 | 2,847.83 | 11,146.10 | 14,564.13 |
| East Nusa Tenggara | 1,452.09 | 2,648.02 | 11,682.02 | 15,030.65 |
| West Kalimantan | 86.87 | 1,196.10 | 6,615.18 | 11,858.41 |
| Central Kalimantan | 21.44 | 295.24 | 1,632.86 | 2,927.07 |
| South Kalimantan | 822.03 | 1,499.05 | 6,613.21 | 8,508.87 |
| East Kalimantan | 20.89 | 287.67 | 1,590.99 | 2,852.02 |
| North Kalimantan | 8.61 | 118.60 | 655.94 | 1,175.83 |
| North Sulawesi | 359.33 | 655.27 | 2,890.81 | 3,719.45 |
| Central Sulawesi | 70.01 | 963.94 | 5,331.22 | 9,556.77 |
| South Sulawesi | 1,311.80 | 2,392.20 | 10,553.42 | 13,578.53 |
| Southeast Sulawesi | 42.89 | 590.48 | 3,265.72 | 5,854.15 |
| Gorontalo | 253.50 | 462.28 | 2,039.40 | 2,623.99 |
| West Sulawesi | 34.82 | 479.45 | 2,651.65 | 4,753.37 |
| Maluku | 11.73 | 161.50 | 893.19 | 1,601.13 |
| North Maluku | 21.08 | 290.19 | 1,604.95 | 2,877.04 |
| West Papua | 411.01 | 542.87 | 2,124.73 | 2,776.29 |
| Southwest Papua | 5,424.02 | 7,164.08 | 28,039.42 | 36,637.89 |
| Papua | 1,954.00 | 2,580.85 | 10,101.16 | 13,198.74 |
| South Papua | 1,212.83 | 1,601.91 | 6,269.68 | 8,192.32 |
| Central Papua | 7,095.03 | 9,371.15 | 36,677.65 | 47,925.10 |
| Highland Papua | 12,653.81 | 16,713.21 | 65,413.70 | 85,473.25 |
| **Indonesia** | **83,162.85** | **119,163.48** | **483,138.35** | **644,215.23** |

Source : Authors' calculation

## Discussion

We have shown the total and unit cost to provide HPV vaccination for OOS girls. The yearly total financial cost reached around US$ 483,000 (1.76% of the national immunization budget allocation in 2025) or US$ 28.92 per fully vaccinated OOS girl, and more than 80% of the total cost are spent on the cost of vaccinating OOS girls aged 11 years. The cost driver at all sites is labor cost and different cities’ characteristics result in different approaches, which in turn results in different total and unit costs. Our results have led to the following observations.

First, our findings suggest that differences in unit costs across study sites were influenced by variation in local implementation context. In Bekasi, the highest unit cost per successfully vaccinated OOS girl was accompanied by labor-intensive target identification and verification processes. Qualitative findings indicated that non-formal institutions were often unstable or no longer operational, requiring repeated physical verification and extensive data matching, which increased time and personnel inputs. Additional operational costs were reported due to security considerations when accessing marginalized populations, involving multi-sectoral teams and higher travel allowances further contributed to program costs. In addition, Bekasi city has the largest population, almost double the population of Bandar Lampung and Bangka city, which may explain the larger unit cost to perform the outreach and vaccination program as it requires extra activities. The approach in Bekasi city requires more coordination and socialization with more stakeholders. Despite these additional operational efforts, the number of vaccinated OOS girls in Bekasi remained lower than in Bandar Lampung, suggesting that greater resource input did not necessarily translate into higher program reach. Local context plays a significant role in this case. Indeed, if we stripped the activities in Bekasi city and only calculated bare minimum activities, the unit costs between regions do not differ much, but it may not reflect the real condition in Bekasi city. In Bandar Lampung city, the largest number of vaccinated OOS girls was achieved through intensive coordination and advocacy activities. Informants described extensive cross-sectoral collaboration and partner-supported target confirmation before field deployment. Personnel time was also required to validate institutional data, as some registered non-formal education facilities did not have eligible OOS girls within the target age group. In Bangka city, implementation was characterized by the integration of OOS outreach into existing community-based activities and the use of localized knowledge from village-level health workers and community leaders. Qualitative findings also highlighted the importance of field validation and travel to geographically dispersed areas in identifying eligible targets. Together, these findings suggest that regional differences in unit costs reflect different operational requirements for reaching OOS girls. Urban target identification in Bekasi, coordination-intensive approaches in Bandar Lampung, and locally integrated outreach in Bangka each contributed to the resources required for program implementation.

Second, the yearly total cost of reaching and vaccinating OOS girls nationally requires around US$ 640,000 per year (28.92 per vaccinated OOS girl), or roughly $2.2million (PPP). [27–32]Our financial cost national estimate only constitutes around 1.76% of the national immunization budget allocation in 2025 (approximately US$ 27.5million) [33] and 4.95% of the HPV vaccine budget in 2022 (around US$ 9.7million), so there is a possibility of economic feasibility to cover the amount, although further assessment is needed. A quick calculation of treating 25% of Indonesian OOS girls (based on national target) of cervical cancer using median treatment cost from a study in Vietnam [34], resulted in around US$ 78.4 million (PPP), basically 35 times of our national coverage and vaccination cost calculation. Given this magnitude, national scale up should be highly considered and the current outreach and HPV vaccination of OOS girls should be optimized to result in higher number of OOS girls getting vaccinated.

While the present study primarily highlights the financial and operational aspects of OOS HPV vaccination, findings from an upcoming study [35] indicate that socio-cultural and trust-related factors may substantially influence uptake. Stakeholders reported concerns regarding vaccine safety, lingering distrust associated with previous immunization programs, and questions surrounding halal status, particularly in some communities. These findings suggest that strengthening community engagement, trust-building initiatives, and locally tailored communication strategies may be as important as ensuring adequate financing.

This study is not without limitations. First, our study only includes several areas in Indonesia. While this restricts the generalizability of our micro costing analysis, our scale up cost estimation method has addressed this limitation and try to accommodate the varied characteristics of regions in Indonesia. However, care still needs to be taken in interpreting our results. Second, the qualitative component was intended to provide explanatory context for the costing findings rather than a comprehensive assessment of implementation barriers. Therefore, additional qualitative research across broader settings may be needed to better understand contextual factors influencing OOS HPV vaccination delivery. Nevertheless, integrating qualitative findings strengthened interpretation of the costing results by providing insight into contextual and operational factors influencing implementation.

## Conclusion

Optimizing OOS girls’ outreach and HPV vaccination is essential to achieve sustained full coverage. Our national cost estimate has shown that reaching and vaccinating OOS girls only costs less than 3% of the national immunization budget, so there is a possibility of economic feasibility, although further assessment is needed. Factors such as local context and implementation challenges should be recognized as they may directly influence the cost and hinder program to scale up.

## Data Availability

The data underlying the results presented in this study contain potentially identifying or sensitive information collected from health facility staff and program implementers in Bekasi City, Bandar Lampung City, and Bangka District, Indonesia. Due to ethical restrictions imposed by Universitas Padjadjaran Ethics Committee. Data cannot be made publicity available. The data set used in this study is available from the corresponding author on reasonable request.

## Appendix

**Table 7.** Respondents.

| No. | Province | Institution | Number of Respondent |
| --- | --- | --- | --- |
| 1 | West Java | Province Health Office | 1 |
|  |  | Bekasi City Health Office | 1 |
|  |  | *Puskesmas* | 4 |
|  |  | Community Health Worker (Kader) | 4 |
|  |  | Other stakeholders | 6 |
| 2 | Lampung | Province Health Office | 1 |
|  |  | Bandar Lampung City Health Office | 1 |
|  |  | *Puskesmas* | 4 |
|  |  | Kader | 4 |
|  |  | Other stakeholders | 6 |
| 3 | Bangka Belitung Island | Province Health Office | 1 |
|  |  | Bangka District Health Office | 1 |
|  |  | *Puskesmas* | 4 |
|  |  | Kader | 4 |
|  |  | Other stakeholders | 6 |

**Table 8.**
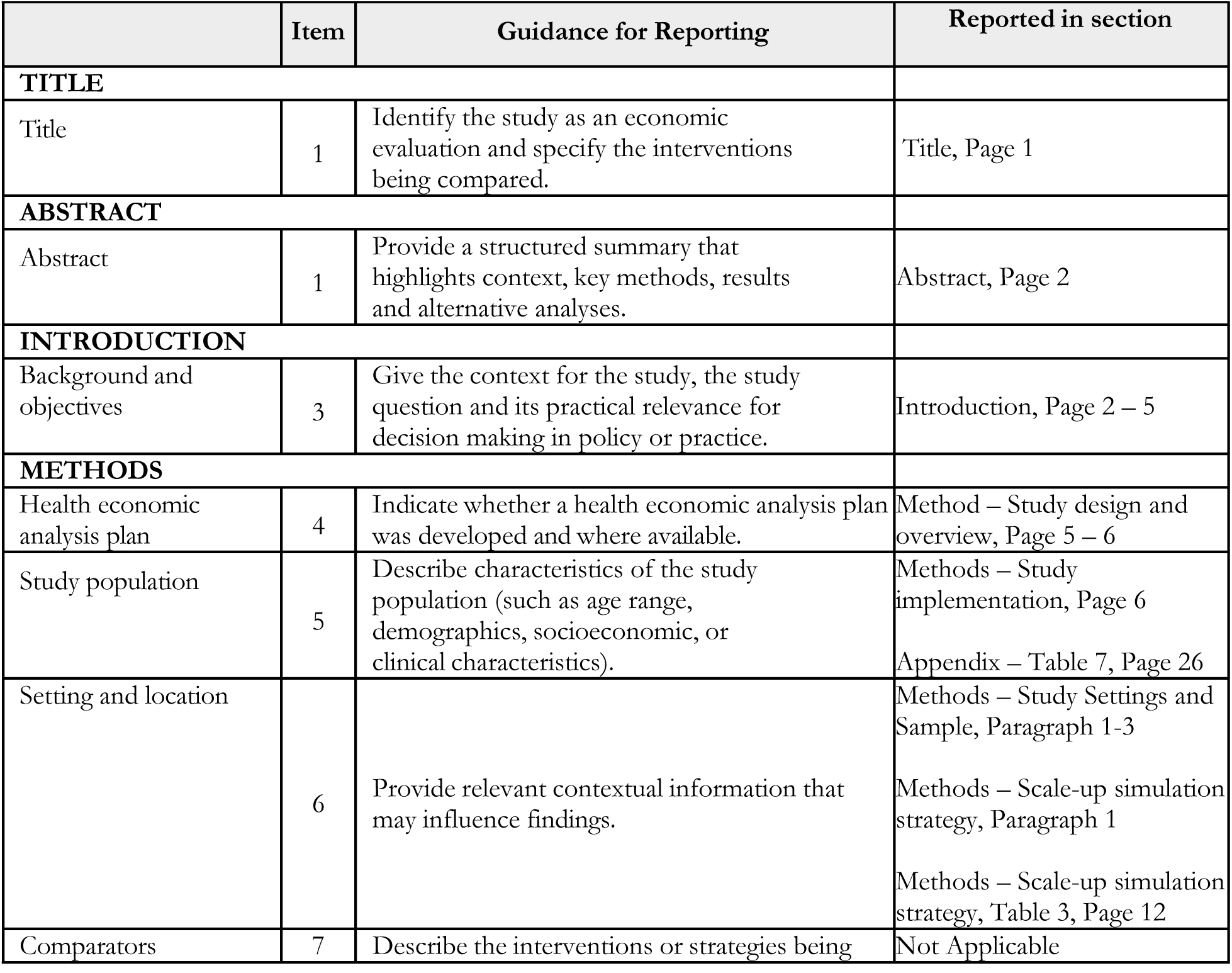
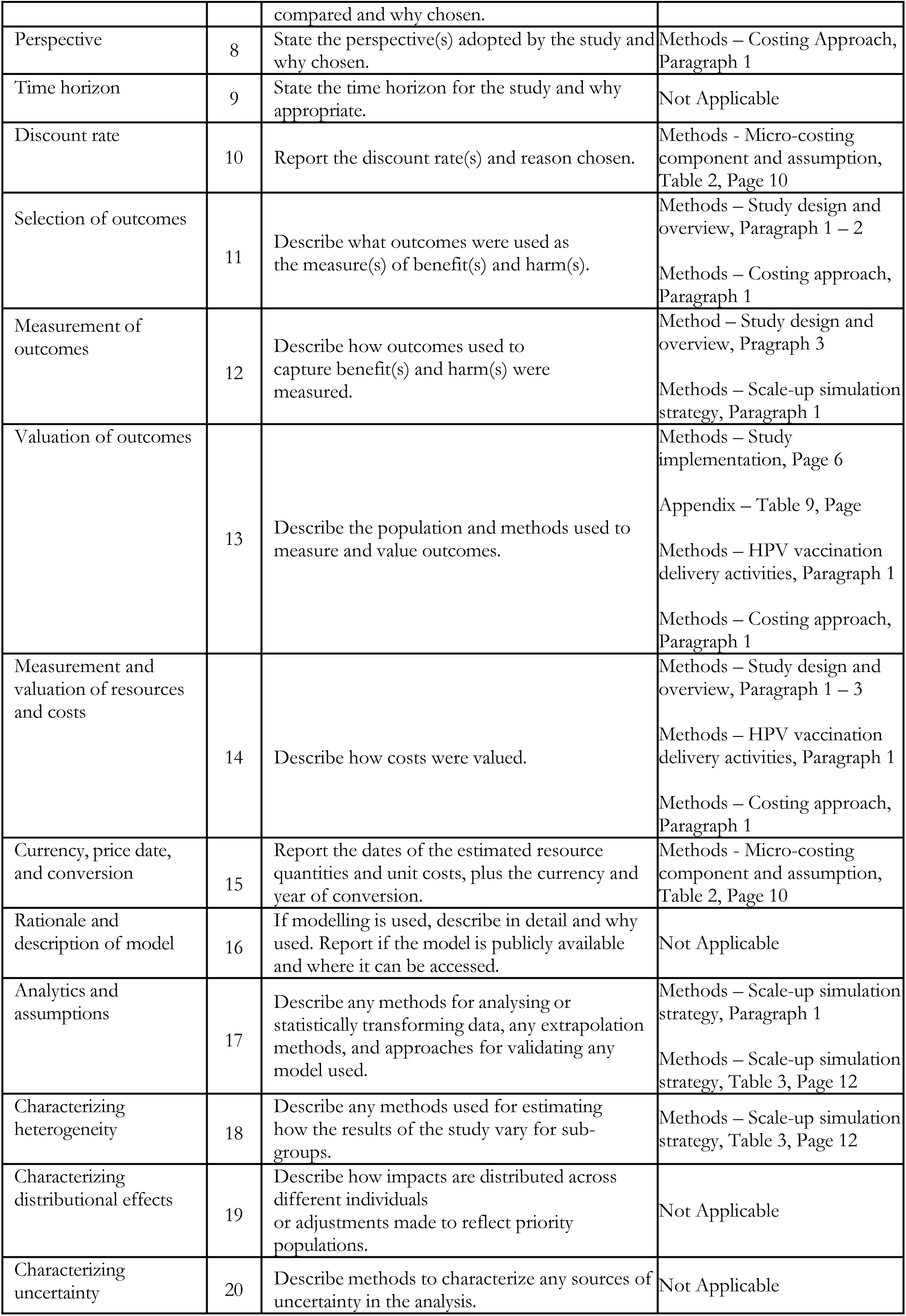
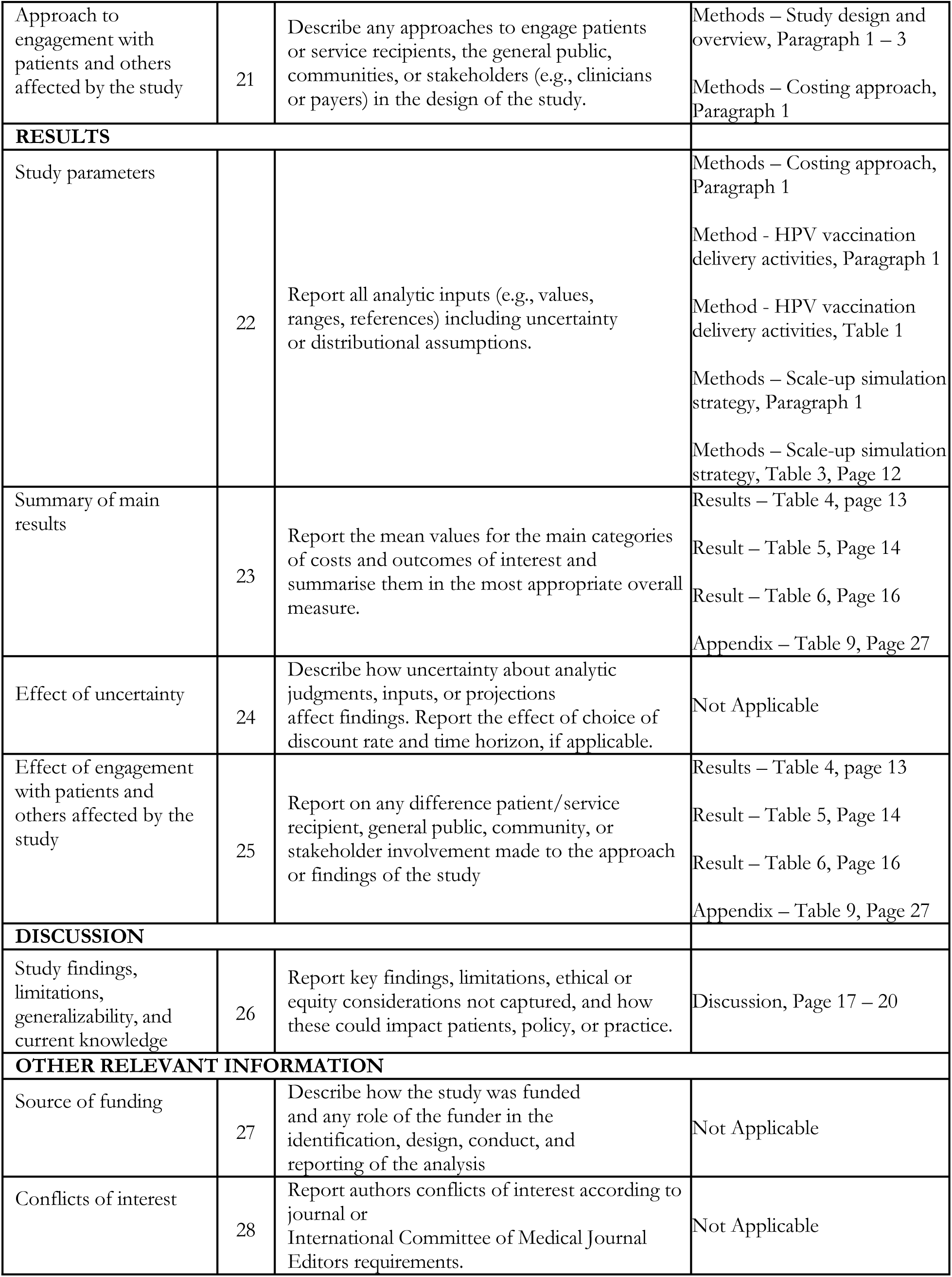
CHEERS 2022 Checklist Report.

**Table 9.** Detailed Assumption of Scale-up Total Cost Estimation.

| Province | Scale-up Mapping | INKINDO 2025 Index | Number of 11 Years old OOS girl |
| --- | --- | --- | --- |
| Aceh | 2 | 0.963 | 294 |
| North Sumatra | 2 | 0.976 | 613 |
| West Sumatra | 2 | 0.947 | 165 |
| Riau | 1 | 1.080 | 154 |
| Jambi | 1 | 0.908 | 168 |
| South Sumatra | 2 | 0.929 | 384 |
| Bengkulu | 2 | 0.889 | 122 |
| Lampung | 2 | 0.899 | 292 |
| Bangka Belitung Islands | 1 | 0.910 | 58 |
| Riau Islands | 2 | 1.101 | 54 |
| Special Capital Region of Jakarta | 3 | 1.000 | 522 |
| West Java | 3 | 0.847 | 1866 |
| Central Java | 3 | 0.823 | 1214 |
| Special Region of Yogyakarta | 3 | 0.838 | 96 |
| East Java | 3 | 0.979 | 2058 |
| Banten | 3 | 0.950 | 492 |
| Bali | 3 | 0.958 | 124 |
| West Nusa Tenggara | 3 | 0.877 | 320 |
| East Nusa Tenggara | 2 | 0.841 | 590 |
| West Kalimantan | 1 | 0.937 | 474 |
| Central Kalimantan | 1 | 0.925 | 117 |
| South Kalimantan | 2 | 0.918 | 334 |
| East Kalimantan | 1 | 1.030 | 114 |
| North Kalimantan | 1 | 1.086 | 47 |
| North Sulawesi | 2 | 0.944 | 146 |
| Central Sulawesi | 1 | 0.908 | 382 |
| South Sulawesi | 2 | 0.970 | 533 |
| Southeast Sulawesi | 1 | 0.947 | 234 |
| Gorontalo | 2 | 0.924 | 103 |
| West Sulawesi | 1 | 0.909 | 190 |
| Maluku | 1 | 0.934 | 64 |
| North Maluku | 1 | 0.940 | 115 |
| West Papua | 3 | 1.202 | 61 |
| Southwest Papua | 3 | 1.194 | 805 |
| Papua | 3 | 1.113 | 290 |
| South Papua | 3 | 1.114 | 180 |
| Central Papua | 3 | 1.130 | 1053 |
| Highland Papua | 3 | 1.153 | 1878 |

Source : Authors' calculation

**Table 10.** Financial Cost: Estimated Cost of HPV Immunization for Out-of-School (OOS) Girls (US$)

| Activities | Bekasi City | Bandar Lampung City | Bangka District |
| --- | --- | --- | --- |
| Target identification, outreach planning, and logistics requirement estimation | 191.84<br>(19.70%) | 21.39<br>(2.15%) | - |
| Target validation: Communication via *whatsapp* | - | 2.26<br>(0.23%) | 1.52<br>(0.92%) |
| Target validation: School visits | 32.27<br>(3.32%) | 52.13<br>(5.23%) | 1.52<br>(0.92%) |
| Socialization with nonformal schools at the *puskesmas* | 165.53<br>(17.00%) | 30.49<br>(3.06%) | - |
| Socialization with nonformal schools: School visits | 30.49<br>(3.13%) | 27.44<br>(2.75%) | - |
| Socialization with nonformal schools: Online | - | - | - |
| Validation of targets from marginalized groups | 30.49<br>(3.13%) | 26.78<br>(2.69%) | - |
| Cost of activities at the district/city office level (table 6) | 0.23<br>(0.02%) | 1.03<br>(0.10%) | 5.96<br>(3.61%) |
| **Total outreach cost** | **450.85**<br>**(46.31%)** | **161.52**<br>**(16.20%)** | **9.01**<br>**(5.45%)** |
| Number of outreach targets | 79.00 | 73.00 | 54.00 |
| *Outreach cost per target* | *5.71* | *2.21* | *0.17* |
| **Total cost of HPV vaccination** | **522.72**<br>**(53.69%)** | **835.29**<br>**(83.80%)** | **156.09**<br>**(94.55%)** |
| Number of targets successfully vaccinated | 33.00 | 56.00 | 13.00 |
| *HPV vaccination cost per target successfully vaccinated* | *20.50* | *17.64* | *13.23* |
| **Total outreach and HPV vaccination cost** | **973.58** | **996.81** | **165.10** |
| *Outreach and HPV vaccination cost per target successfully vaccinated* | *29.50* | *17.80* | *12.70* |

Note : The percentage for each cost component is calculated by dividing the component cost by the total outreach and HPV vaccination cost.
Source : Authors' calculation

